# Rethinking Tobacco Policy: Why a Cap-and-Levy Scheme Can Outperform the UK’s Sales Ban

**DOI:** 10.1101/2025.05.28.25328485

**Authors:** Joan E. Madia

**Affiliations:** Nuffield Deparment of Primary Care Health Sciences, University of Oxford

**Keywords:** Smoke-free, Generational Sales Ban, Cap-and-levy, Illicit tobacco market, Smoking prevalence

## Abstract

The United Kingdom aims to be smoke-free by 2030, and in pursuit of this goal, has proposed a Generational Sales Ban (GSB). While somewhat innovative, the GSB exclusively targets future generations, potentially overlooking the immediate health burdens and illicit market risks associated with current smokers. This study argues for a cap-and-levy scheme as a more comprehensive and efficient alternative. By directly addressing supply, consumption, and state tax revenues, a cap-and-levy approach offers a broader impact on smoking prevalence including existing smokers, while potentially mitigating the unintended consequences of a sales ban, such as fuelling the illicit trade and reducing tax revenues. The provision of accessible and appealing alternatives to combustible tobacco is crucial in minimizing the appeal of illicit products under any restrictive policy. This analysis suggests that a cap-and-levy mechanism warrants consideration as a policy instrument that could outperform the GSB in achieving significant and immediate reductions in smoking-related harm, without the unintended consequences that the GSB would produce.

**Highlights:**

- The UK remains short of the 2030 smoke-free goal despite declining smoking rates.
- The government proposes a Generational Sales Ban (GSB) prohibiting sales to those born after 2009.
- Studies suggest that GSB may fuel the illicit tobacco market (26% of UK consumption) and reduce tax revenues.
- This study advocates a cap-and-levy scheme as a more effective alternative for achieving the UK’s smoke-free ambitions.
- Providing appealing alternatives curbs illicit tobacco use under strict regulations.

## Introduction

Despite steady declines in smoking rates, the United Kingdom remains short of its goal to become smoke-free by 2030, defined as achieving a smoking prevalence below 5% (Department of Health and Social Care, 2019). In response, the government has proposed a Generational Sales Ban (GSB), which prohibits the sale of tobacco to anyone born after 1 January 2009. While innovative, this approach narrowly targets future generations, leaving out existing smokers who continue to account for the majority of tobacco-related harm and healthcare costs (ONS, 2023). Research indicates that smoking prevalence is consistently higher among lower socioeconomic status groups who face greater challenges in quitting (Reid et al., 2010; Hiscock et al., 2012).

Additionally, the GSB may inadvertently fuel the illicit tobacco market - already comprising 26% of UK consumption (HMRC, 2023) - and reduce tax revenues. Recent research by Cotti et al. (2024) on similar age-restriction policies found that treated smokers often adapt by purchasing cigarettes in different jurisdictions rather than quitting entirely. Public smoking bans have shown limited impact on overall smoking prevalence (Jones et al., 2011) and may even produce unintended consequences such as displacing smoking to private spaces (Adda & Cornaglia, 2010), without delivering substantial public health gains in the short to medium term. In this regard, to effectively reduce the appeal of the illicit tobacco market, it is crucial to provide a wide array of attractive substitute products. Expanding the availability and variety of legal alternatives can help transition smokers away from combustible tobacco while diminishing the demand for illicit options. This strategy aims to offer current smokers viable alternatives, making it less likely they will turn to the illicit market due to a lack of appealing choices.

This study addresses the urgent need for a more effective and economically sustainable strategy to phase out combustible tobacco use in the UK. By evaluating a proposed Cap-and-Levy Scheme, which would progressively reduce the legal supply of tobacco while introducing a higher tax on excess production, the analysis offers a compelling alternative to the GSB. Drawing inspiration from environmental policy tools like the EU Emissions Trading System and the UK Vehicle Emissions Trading Scheme (World Bank, 1999), and responding to evidence that higher tobacco taxes are most effective in reducing tobacco exposure (Adda & Cornaglia, 2010), the cap-and-levy model combines supply restriction with economic incentives to encourage cessation and promote healthier alternatives, while addressing concerns about the regressive impact of tobacco taxes on low-income households (Darden et al., 2025) through targeted cessation support (Brown et al., 2014).

This study makes a twofold contribution. First, it critically evaluates the limitations of the UK’s proposed Generational Sales Ban (GSB), showing that it is unlikely to deliver the country’s smoke-free target due to its narrow focus on future cohorts and minimal short-to medium-term public health gains. In contrast, the cap-and-levy scheme emerges as a more effective and fiscally sustainable alternative. Using projections on smoking prevalence, illicit trade, and tax revenue, we show that the cap-and-levy could reduce smoking below the 5% threshold by 2039 while generating £14.3 billion more in public revenue than existing policy trajectories (HMRC, 2023; Public Health England, 2018). Second, this work demonstrates that the cap-and-levy approach balances public health and economic goals more effectively than the GSB, offering a practical path to phasing out combustible tobacco.

## Background and Literature Review

Despite steady declines in smoking rates, the United Kingdom remains short of its goal to become smoke-free by 2030, defined as achieving a smoking prevalence below 5% (Department of Health and Social Care, 2019). In response, the government has proposed a Generational Sales Ban (GSB), which prohibits the sale of tobacco to anyone born after 1 January 2009. While innovative, this approach narrowly targets future generations, leaving out existing smokers who continue to account for the majority of tobacco-related harm and healthcare costs (ONS, 2023).

### Tobacco Control Policies and Their Effectiveness

Research on generational tobacco bans and smoking cessation interventions reveals mixed outcomes for addressing existing smokers’ needs. Critiques of these bans highlight challenges such as the risk of increased black market activity and limited short-term efficacy in reducing smoking rates (Snowdon, 2024). Evidence indicates that smoking prevalence has been declining over time, supported by broader social changes such as the rise of home and workplace smoking bans (Levy et al., 2004). Contrary to the “hardening” hypothesis - which suggests that the remaining smoking population becomes increasingly resistant to quitting - recent findings support a “softening” trend: as prevalence declines, smokers exhibit higher quit attempts and lower cigarette consumption (Kulik & Glantz, 2016).

Recent research on Tobacco 21 (T21) laws, which raise the minimum legal sales age to 21, offers insights into age-restriction approaches similar to the proposed GSB. Cotti et al. (2024) found that T21 laws reduce self-reported cigarette smoking among 18-to-20-year-olds, with effects concentrated among males. Their research also revealed that individuals who “age-out” of treatment are less likely to subsequently initiate smoking or vaping, suggesting potential long-term benefits of age-restriction policies. However, behavioural adaptations were observed, with treated smokers less likely to buy their own cigarettes and more likely to purchase cigarettes in different states. Importantly, the authors found evidence of non-classical measurement error, with T21 laws reducing the probability that clinically identified likely smokers self-report as smokers -suggesting that self-reported reductions may overstate actual behaviour changes.

The impact of smoking bans on overall smoking prevalence appears limited, though some effects on consumption levels among specific groups have been observed (Jones et al., 2011). Studies of smoking bans in healthcare facilities have shown no major negative impacts on behaviour but little effect on cessation rates (el-Guebaly et al., 2002). Paradoxically, public smoking bans may increase non-smokers’ exposure to tobacco smoke by displacing smokers to private spaces, particularly affecting children (Adda & Cornaglia, 2010). In contrast, progressively increased tobacco taxes have been found to be more effective in reducing exposure to tobacco smoke, especially for those most at risk (Adda & Cornaglia, 2010).

### Socioeconomic Disparities in Smoking

A significant challenge in tobacco control is addressing socioeconomic disparities in smoking behavior. Smoking prevalence is consistently higher among lower socioeconomic status (SES) groups across Western countries, with these individuals facing greater challenges in quitting (Reid et al., 2010; Hiscock et al., 2012). Individual-level cessation interventions in Europe may have reduced overall smoking rates but often increased inequalities, with lower SES groups less likely to quit. Multiple factors contribute to lower cessation rates among disadvantaged smokers, including age, proportion of smoking friends and family, nicotine dependence, and medication use (Hiscock et al., 2015).

Low SES smokers tend to have more smoking friends and are more likely to gain smoking friends over time, potentially reinforcing their smoking behaviour (Hitchman et al., 2014). This social network effect creates additional barriers to cessation for this population. However, targeted UK National Health Service stop-smoking services have shown positive equity impacts by achieving higher uptake among low-SES smokers, potentially compensating for lower quit rates in this group (Brown et al., 2014). These findings suggest that targeted interventions may be more effective in addressing cessation needs across diverse populations.

To combat high smoking rates in disadvantaged groups, a comprehensive approach combining various tobacco control measures is necessary, alongside broader efforts to address health inequalities (Hiscock et al., 2012). Among potential interventions, raising tobacco prices appears most effective in reducing health inequalities, though targeted cessation programs and mass media campaigns may also help (Hiscock et al., 2012).

### Economic Impacts and Unintended Consequences

Research on smoking bans reveals complex economic impacts and unintended consequences that must be considered when designing tobacco control policies. While some studies claim bans don’t harm businesses, evidence suggests differential effects across the hospitality industry (Craven & Marlow, 2008; Marlow, 2010). Restaurants often benefit or remain unaffected, while bars and fraternal organizations may experience losses, as indicated by noncompliance patterns (Marlow, 2010). Aggregate data analysis can mask these distributional effects, overlooking important economic inefficiencies (Pakko, 2006).

Recent research by Darden et al. (2025) on cigarette taxes reveals concerning unintended consequences for household budgets, particularly among lower-income populations. Their study found that while cigarette taxes reduce smoking prevalence, they increase cigarette expenditures among continuing smokers. More troublingly, a $1 increase in cigarette taxes caused a 2.12% decline in human capital-related expenditures among below-median income smokers, suggesting that tax policies may inadvertently reduce spending on education, health, and other essential services for vulnerable populations. Their experimental survey also showed that smokers respond to tax increases by adjusting cigarette shopping behaviours, substituting toward other tobacco products, and reducing both discretionary and essential expenditures.

Other unintended consequences of bans may include increased drunk driving as patrons travel further to smoke and drink, and reduced innovation in air filtration technology (Marlow, 2009). The Coasian framework suggests that private markets can address smoking disputes without government intervention (Craven & Marlow, 2008). These economic considerations are particularly relevant to the GSB, which may inadvertently fuel the illicit tobacco market - already comprising 26% of UK consumption (HMRC, 2023) - and reduce tax revenues, without delivering substantial public health gains in the short to medium term.

### The Case for a Cap-and-Levy Approach

Given the limitations of traditional smoking bans and the GSB, this study addresses the urgent need for a more effective and economically sustainable strategy to phase out combustible tobacco use in the UK. By evaluating a proposed Cap-and-Levy Scheme, which would progressively reduce the legal supply of tobacco while introducing a higher tax on excess production, the analysis offers a compelling alternative to the GSB.

Drawing inspiration from environmental policy tools like the EU Emissions Trading System and the UK Vehicle Emissions Trading Scheme (World Bank, 1999), the cap-and-levy model combines supply restriction with economic incentives to encourage cessation and promote healthier alternatives. This approach addresses the socioeconomic disparities highlighted in the literature by using taxation, which has proven effective for disadvantaged populations, while simultaneously targeting supply reduction to accelerate progress toward a smoke-free UK.

## Theoretical Framework

### Cap-and-Levy as a Regulatory Innovation in Tobacco Control

Tobacco control policies have historically relied on demand-side interventions such as public education, taxation, and age-based restrictions. However, the persistence of smoking— particularly among older and socioeconomically disadvantaged populations—has highlighted the limits of these approaches in achieving rapid, population-wide declines in tobacco use. In this context, the proposed cap-and-levy phase-out scheme offers a novel supply-side strategy, targeting both the availability and affordability of combustible tobacco products in a progressively structured manner.

The cap-and-levy approach draws from environmental economics and regulatory design. Similar frameworks have proven effective in non-health domains, such as the EU Emissions Trading System (EU ETS) and the UK Vehicle Emissions Trading Scheme, where market-based caps on harmful outputs incentivize reductions and innovation without requiring full bans. Applying this logic to tobacco regulation allows policymakers to gradually phase out smoking while minimizing unintended consequences like illicit trade or consumer backlash.

Under the proposed scheme, beginning in 2027, the government would impose an annual cap on the total volume of combustible tobacco products that can be legally sold in the UK. The initial cap would be set at 85% of the previous year’s sales volume, with annual reductions implemented at a fixed rate. This means the cap is reduced by 15% each year relative to the previous year’s limit, not cumulatively—i.e., the cap follows a compounding decline rather than a linear one. This cap would be divided into tradable allowances allocated to tobacco companies in proportion to their historical market share. To introduce price disincentives, companies wishing to sell up to 4% beyond their allowance could do so by paying an additional levy formed by a specific levy increased by 40% (Figure 1). With the cap-and-levy approach, the volume would fall to half the 2027 level by 2035, compared to 2038 under the business-as-usual (BAU) scenario.

**Figure 1:**
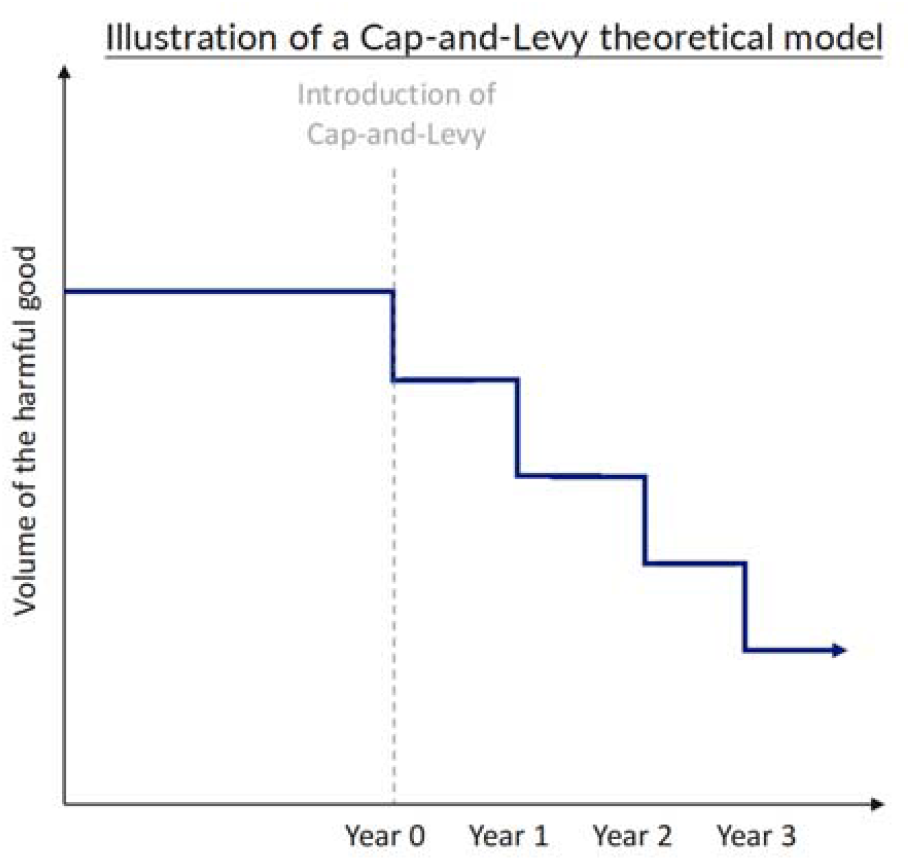
Theoretical impact of a cap-and-levy scheme on product supply. *Notes: A stylized representation of how a gradually decreasing cap, combined with a fiscal levy on over-cap sales, restricts product availability while maintaining regulatory flexibility*.

This dual mechanism - restricting supply and raising prices - creates mutually reinforcing incentives to reduce consumption and encourages both producers and consumers to transition to lower-risk alternatives (e.g., e-cigarettes or heated tobacco products). Importantly, by regulating producers rather than end-users, the cap-and-levy simplifies enforcement and mitigates the regulatory burdens associated with age verification and retail surveillance, which have historically limited the effectiveness of age-based bans.

By explicitly incorporating behavioural economics and regulatory flexibility into its design, the cap-and-levy model represents a paradigm shift in tobacco control - moving from reactive deterrence to proactive phase-out. It is especially relevant in high-income countries like the UK, where baseline smoking rates are already low but where further reductions require addressing entrenched consumer segments and overcoming diminishing marginal returns from traditional policies.

## Methods and Data Sources

### Simulation Approach

This analysis evaluates the projected health and fiscal outcomes of two regulatory strategies aimed at reducing combustible tobacco use in the United Kingdom: the Generational Sales Ban (GSB) and a Cap-and-Levy phase-out scheme. Both are compared to a business-as-usual (BAU) scenario using a simulation model built in Excel. The model estimates smoking prevalence, legal tobacco sales volumes (in billion stick equivalents), tax revenue from both combustible and non-combustible products, and the share of illicit tobacco use over time, with a projection horizon running from 2021 to 2040.

The Cap-and-Levy approach represents a modern economic tool designed to phase out harmful products through regulated supply restriction. Unlike traditional demand-side interventions, this supply-side mechanism establishes a declining cap on combustible tobacco products while allowing manufacturers limited flexibility through a levy system. This approach draws inspiration from established phase-out schemes such as the EU Emissions Trading System, vehicle emissions trading schemes in the UK, and proposed transferable exclusivity vouchers for pharmaceuticals.

### Policy Scenario Designs

The GSB scenario assumes a ban on sales to individuals born after 1 January 2009, beginning in 2027. Its impact is limited by design to future cohorts, with no effect on the current smoking population. The model assumes a 33% reduction in smoking initiation probability for those affected by the ban, based on previous tobacco control policy evaluations.

The Cap-and-Levy scheme, by contrast, introduces a national cap on the legal supply of combustible products, starting at 85% of the previous year’s volume and decreasing by 15% annually. Companies are allocated sales allowances proportional to their average market share over the preceding five years. Two distinct scenarios were developed for this approach:

#### The Cap-and-Levy Efficiency-Oriented Scenario

This scenario prioritizes fiscal sustainability while still contributing to public health goals. It aims to demonstrate that environmental and health objectives can be achieved without significant government revenue loss, potentially making the policy more politically viable in resource-constrained environments. Companies may sell up to an additional 8% of volume under an increased (+75%) specific tax. The ad valorem component remains the same (16.5%). This creates enough punishment for market players to have to deeply think if they want to move in this direction. On the plus side, ensures also substantial revenue generation.

#### The Cap-and-Levy Effectiveness-Oriented Scenario

This scenario prioritizes public health outcomes and accelerated achievement of the UK’s smoke-free goal, accepting a fiscal trade-off. It demonstrates how the same policy framework can be calibrated to maximize health benefits when revenue considerations are of secondary importance. Companies may sell up to an additional 4% of volume under a higher specific tax rate (+40%). The ad valorem component remains the same (16.5%). While the specific tax increase is lower than in the efficiency-oriented scenario, market players have a smaller room for increasing their sales above the cap (only 4% vs 7%). This results in better health results. The clear trade-off is tax revenues, which are considerably reduced in this scenario.

### Model Assumptions and Parameters

All scenarios assume a 5% shift to the illicit market due to regulatory tightening. However, the model also assumes that 90% of smokers who quit under the Cap-and-Levy scenario will switch to taxed non-combustible alternatives (e.g., heated tobacco products or e-cigarettes), partially offsetting losses in combustible tax revenues. This shows the importance of alternatives for any scenario.

The model’s baseline assumptions include no dual use between legal and illicit products, equivalent consumption patterns between legal and illicit smokers, illicit market prices following inflation, and implementation of both policy interventions beginning in 2027.

Age-cohort analysis segments smoking prevalence across age groups (14-30, 31-54, 55+) to capture differential impacts across the population, particularly important given the age-specific targeting of the GSB policy. Fiscal projections incorporate tax revenue estimates for both combustible products and potential revenue from non-combustible alternatives through 2040.

### Data Sources and Elasticity Modelling

The model incorporates historical data sources from the UK Government Tobacco Bulletin, Office for National Statistics population projections, and smoking prevalence surveys from 2017-2023. The model applies fixed price elasticity of demand values derived from the empirical literature. These include estimates from Pryce et al. (2023), based on UK data from 2006 to 2017, and Prieger and Kulick (2016), which offer complementary EU-level estimates across factory-made cigarettes (FMC), fine-cut tobacco (FC), and non-duty-paid (NDP) tobacco categories. This matrix of elasticities allows for cross-product substitution effects to be captured when prices change under different policy scenarios.

Illicit market prices were calculated using a weighted average of cigarette prices in countries identified as major sources of illicit supply. For example, the price of the most-sold brand in Poland, Romania, and Turkey was used to estimate an average illicit price per pack in the UK, with weights reflecting each country’s contribution to the total illicit inflow. This allowed the model to estimate switching behaviour more realistically under price shocks from either policy scenario. A list of the primary data sources used to construct the model is provided in Table 1, while full details on the data, assumptions, and modelling approaches are available in the Appendix.

**Table 1.**
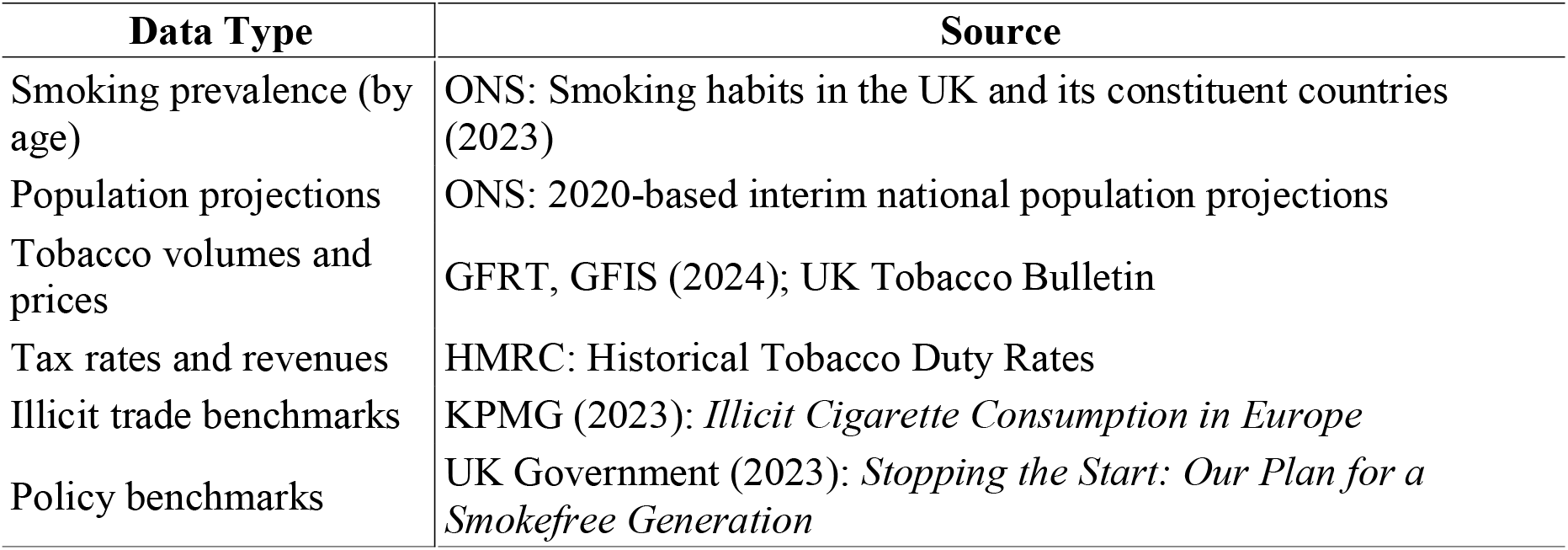
Primary data sources used in the simulation model.

While the analysis incorporates certain simplifying assumptions, including the use of static elasticity estimates, the potential for regional variations in illicit market prices, the homogeneous treatment of smoking rates within age groups, and assumptions regarding switching behaviour, these do not undermine the study’s central findings. Sensitivity analyses have been conducted to test the model’s robustness, and the results consistently affirm the comparative advantages of the Cap-and-Levy approach over the Generational Sales Ban (GSB). This methodological framework provides a valuable tool for demonstrating how the Cap-and-Levy mechanism offers policymakers greater flexibility in balancing public health and fiscal objectives, with the potential to accelerate progress toward smoke-free goals.

## Results

### Generational Sales Ban

The UK’s current strategy to become smoke-free by 2030 - a target defined as achieving smoking prevalence below 5% - has included the proposed Generational Sales Ban (GSB), which prohibits the sale of tobacco products to anyone born on or after 1 January 2009. While this policy may accelerate the long-term decline in smoking prevalence, it is unlikely to achieve the smoke-free target within the desired timeframe due to its narrow focus. By 2030, the GSB would only affect 6% of potential smokers, increasing to just 22% by 2040. Projections show that smoking prevalence under the GSB would fall to 8.9% by 2030 and 5.8% by 2040, a mere 0.3 percentage point lower than under a business-as-usual (BAU) scenario (Figure 2).

**Figure 2:**
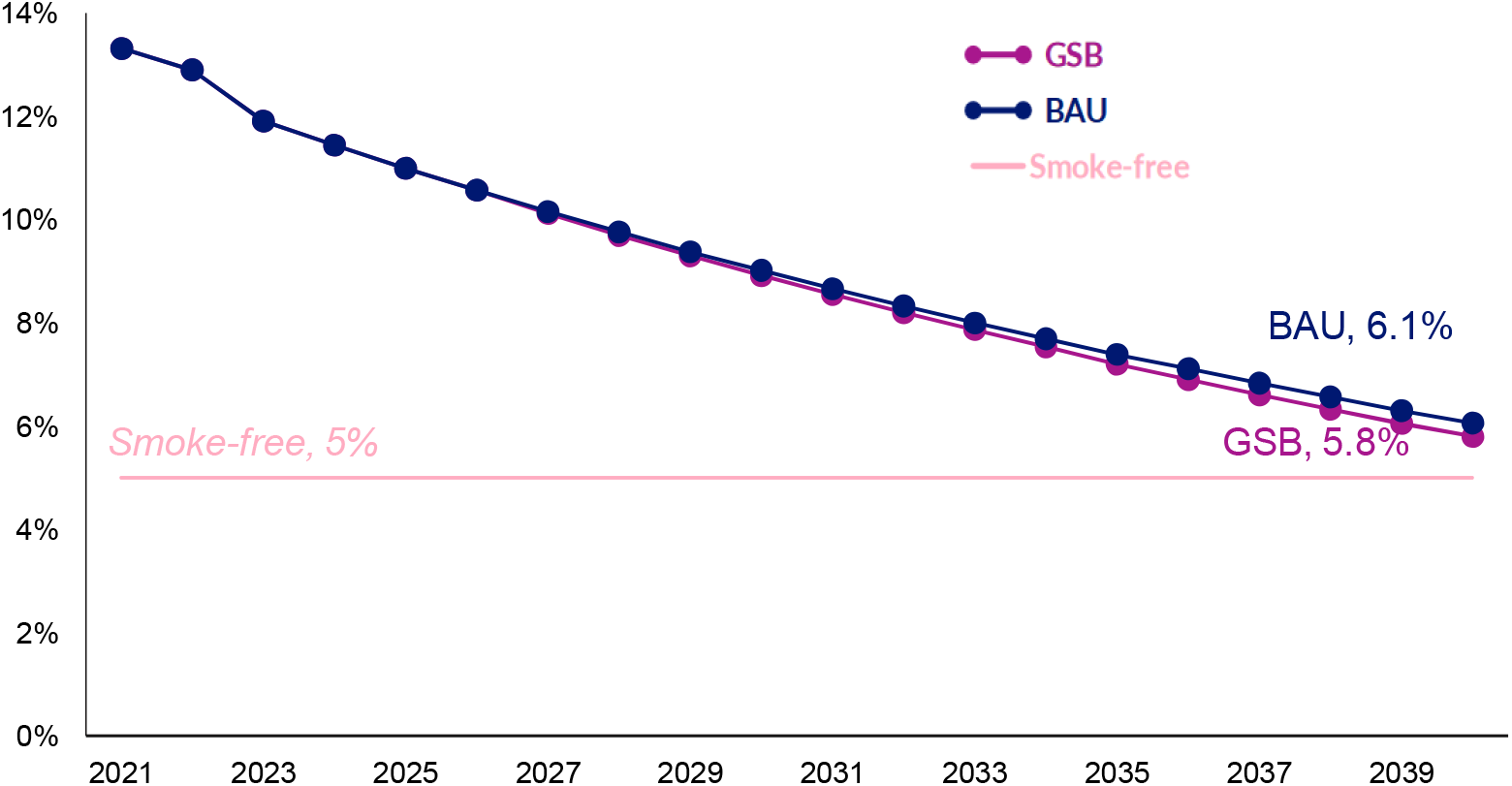
Projected impact of a Generational Sales Ban on smoking prevalence, 2023– 2040. *Notes: The GSB leads to modest reductions in smoking prevalence relative to BAU, with limited population coverage by 2040. Among 18+ population*.

**Figure 2:**
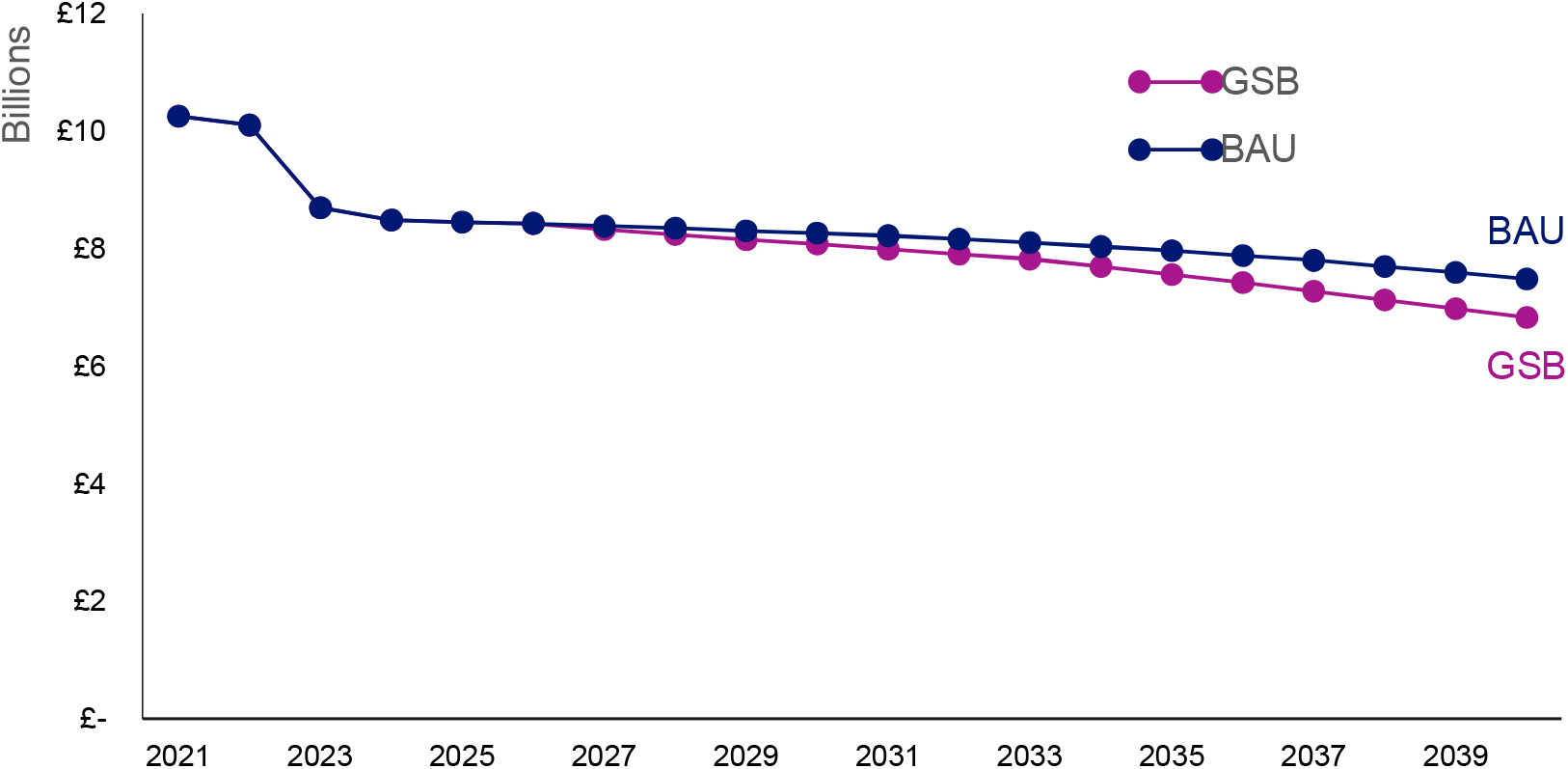
Projected tax revenue under the Generational Sales Ban, 2023–2040. *Notes: Total tax revenue from legal tobacco sales declines by £4*.*9 billion compared to the BAU scenario*.

Furthermore, the policy disproportionately targets youth, whose smoking rates have already declined sharply - from 24% in 2018 to 8% in 2023 - while having little effect on older cohorts, among whom smoking rates have declined marginally or even increased (e.g., +0.3% among those aged 65+). This demographic dynamic undermines the policy’s potential public health benefits and prolongs pressure on the NHS.

As illustrated in Figure 3, the GSB’s demographic focus critically limits its ability to produce meaningful reductions across the population. While smoking among younger demographics has already seen remarkable declines - dropping from 24% in 2018 to 8% in 2023 - the policy paradoxically concentrates its efforts on this very group. In contrast, older cohorts, who are responsible for the greatest share of tobacco-related morbidity and NHS burden, are effectively overlooked.

**Figure 3:**
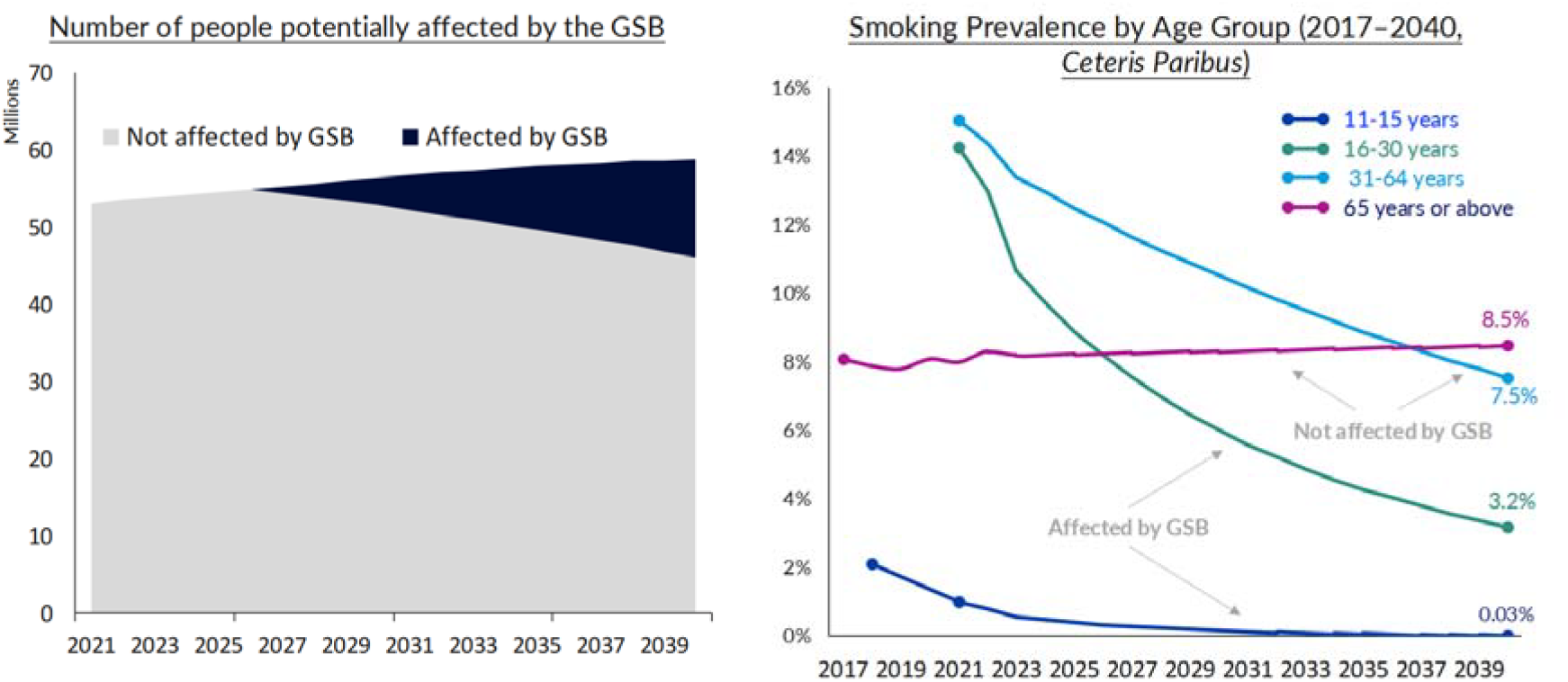
Demographic Limitations of the Generational Sales Ban. *Notes: Left Panel: Number of people potentially affected by the GSB from 2021 to 2039, showing the limited population coverage. Right Panel: Smoking prevalence by age group from 2017 to 2040, highlighting the uneven impact of the GSB across demographic cohorts*.

**Figure 3.**
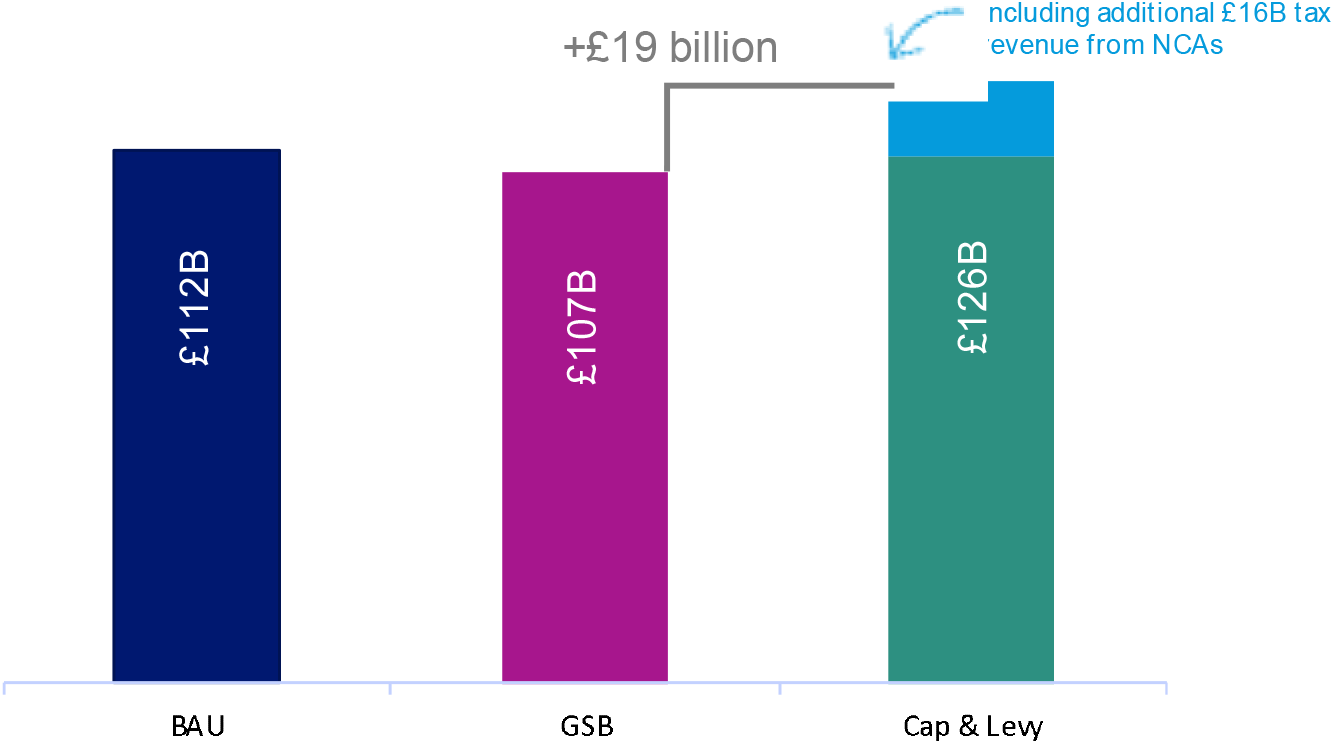
Projected tax revenue under the Cap-and-Levy Scheme, 2027–2040. *Notes: The cap-and-levy model increases cumulative tax revenues by £14*.*3 billion vs. BAU and £19*.*2 billion vs. GSB*. Smokers switching to non-combustibles will add £16.0bn in tax revenue. Assuming 90% switching to NCP.

This uneven targeting is particularly concerning given the stagnation in smoking prevalence among older groups. The 55–64 age group has seen only a marginal decline of 1.3%, while the 65+ age group has experienced a slight increase of 0.3%. These older populations face the highest risks of smoking-related health complications and generate the most substantial healthcare costs, yet the GSB offers them no meaningful pathway to cessation.

By design, the GSB essentially writes off current smokers - particularly those in older age brackets - creating a policy approach that is both demographically myopic and potentially discriminatory. It establishes a two-tier system of tobacco control, where younger generations are subject to comprehensive intervention, while older smokers are left behind. This not only limits the policy’s effectiveness but also risks perpetuating health inequalities by providing differential support based on arbitrary age boundaries.

### Efficiency Scenario Results

The Efficiency scenario for the Cap-and-Levy scheme aims to maintain tax revenues at levels comparable to the Generational Sales Ban (GSB) while effectively reducing smoking prevalence. Under this scenario, a 15% cap on combustible products is introduced in 2027 with annual reductions, complemented by allowing firms to sell 7% more volume by paying an increased (+75%) specific tax (levy).

From a fiscal perspective, the GSB is projected to reduce tax revenues by £4.9 billion by 2040 (Figure 2). This substantial decline results primarily from its impact on legal sales, coupled with an anticipated increase in illicit consumption, which already constitutes 26% of the UK market. These unintended consequences could significantly erode public funding for essential services, including healthcare.

In contrast, the cap-and-levy scheme generates considerably higher fiscal returns. Between 2027 and 2040, it is expected to yield £14.3 billion more in tax revenue than the BAU scenario and £19.2 billion more than the GSB (Figure 3) that includes £16.0 billion contribution from non-combustible products. This enhanced revenue stream reflects the scheme’s ability to maintain and expand the tax base through higher levy rates on permitted over-cap sales, while simultaneously reducing smoking prevalence to the level of the GSB.

This dual impact - effectively reducing smoking while increasing revenue - positions the cap-and-levy as a dominant strategy over the GSB from both public health and public finance perspectives.

Figure 4 illustrates the comprehensive outcomes of this scenario. The Cap-and-Levy scheme achieves a smoking prevalence of 5.8% by 2040, matching the GSB scenario and significantly lower than the 6.1% projected in the business-as-usual (BAU) scenario. Importantly, unlike the GSB which primarily benefits younger individuals, the Cap-and-Levy reduces smoking across all age groups.

**Figure 4:**
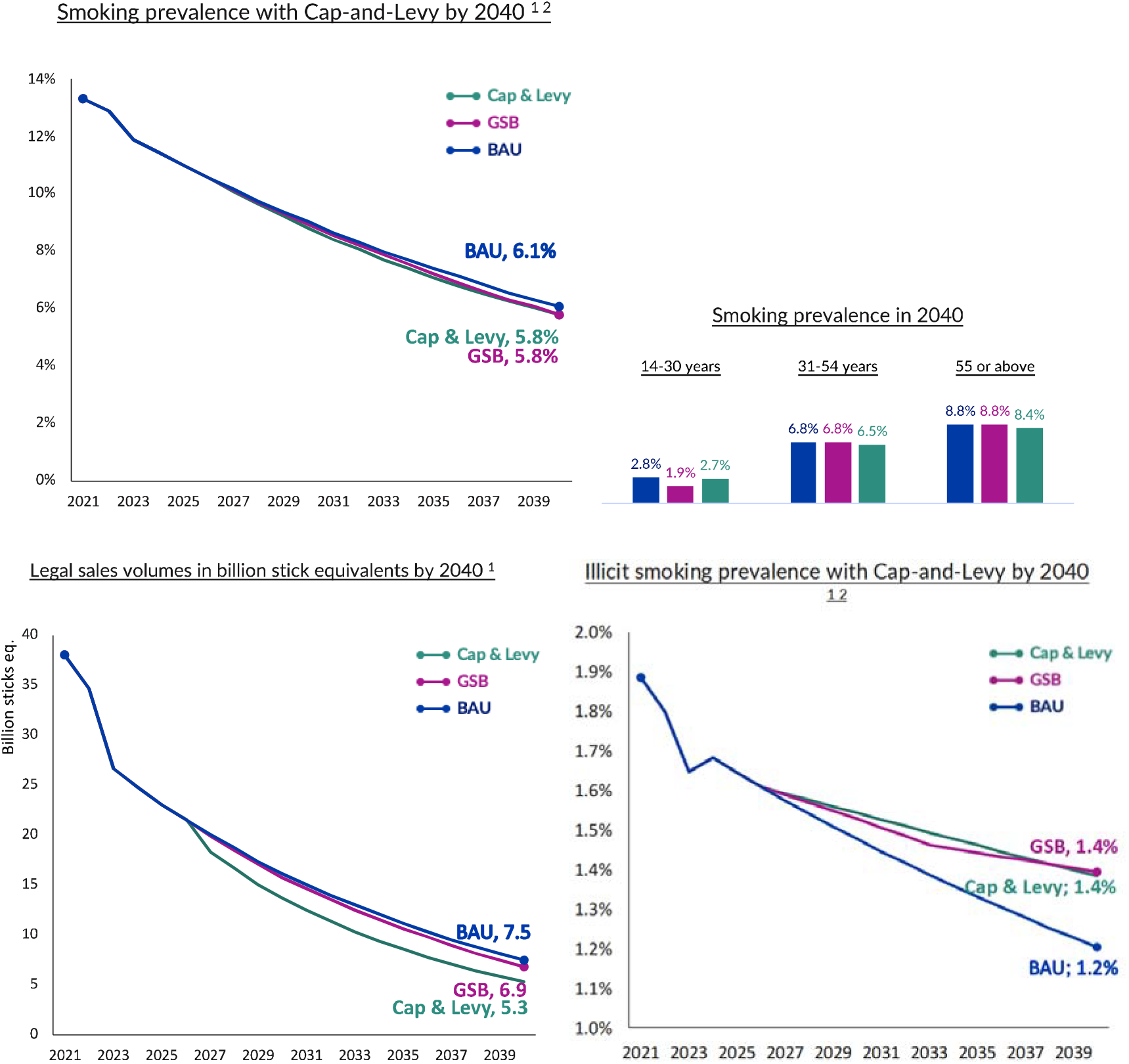
Smoking prevalence, Legal sales volume, and Illicit Smoking, under Efficiency Scenario.

The Cap-and-Levy scheme also drives a faster decline in legal sales volumes compared to BAU, though slightly slower than the GSB. By 2040, legal sales volumes reach 5.7 billion stick equivalents under Cap-and-Levy, compared to 6.9 billion under GSB and 7.5 billion under BAU. This reduction stems from the cap limiting supply and the levy increasing prices, both contributing to lower consumption.

Both the GSB and Cap-and-Levy scenarios lead to an increase in illicit trade due to tighter restrictions on the legal market, with illicit smoking prevalence reaching 1.4% by 2040 under both scenarios, compared to 1.2% under BAU.

In summary, while the generational sales ban (GSB) offers a symbolic step toward reducing youth smoking, its practical impact on smoking prevalence, public health burden, and fiscal outcomes remains limited in the medium term. By contrast, the cap-and-levy scheme offers a more comprehensive and enforceable policy tool that targets current and future smokers alike. It achieves comparable reductions in smoking prevalence by 2040 while significantly increasing government revenue and minimizing incentives for illicit trade. These dual gains - improved health outcomes and stronger fiscal performance-position the cap-and-levy scheme as a superior alternative to the GSB. This finding is particularly relevant given strained healthcare resources and the need for sustainable policy solutions that align health and economic incentives.

### Effectiveness Scenario Results

The “Effectiveness” scenario prioritizes the elimination of combustible consumption across all age groups. In this scenario, the cap is set at 15% with annual reductions from 2027, and firms can sell an additional 4% of volume by paying an the increased (+40%) specific tax (levy).

In contrast to the GSB, a cap-and-levy scheme -consisting of a progressively decreasing cap on tobacco supply and a levy imposing higher ad valorem taxes on excess volumes - demonstrates superior effectiveness. Projections indicate that cap-and-levy could reduce smoking prevalence to 4.7% by 2040, achieving the smoke-free goal by 2039 (Figure 5). This outcome reflects the broader population reach of the cap-and-levy approach and its dual supply-demand pressure: limiting availability while raising prices to discourage consumption.

**Figure 5:**
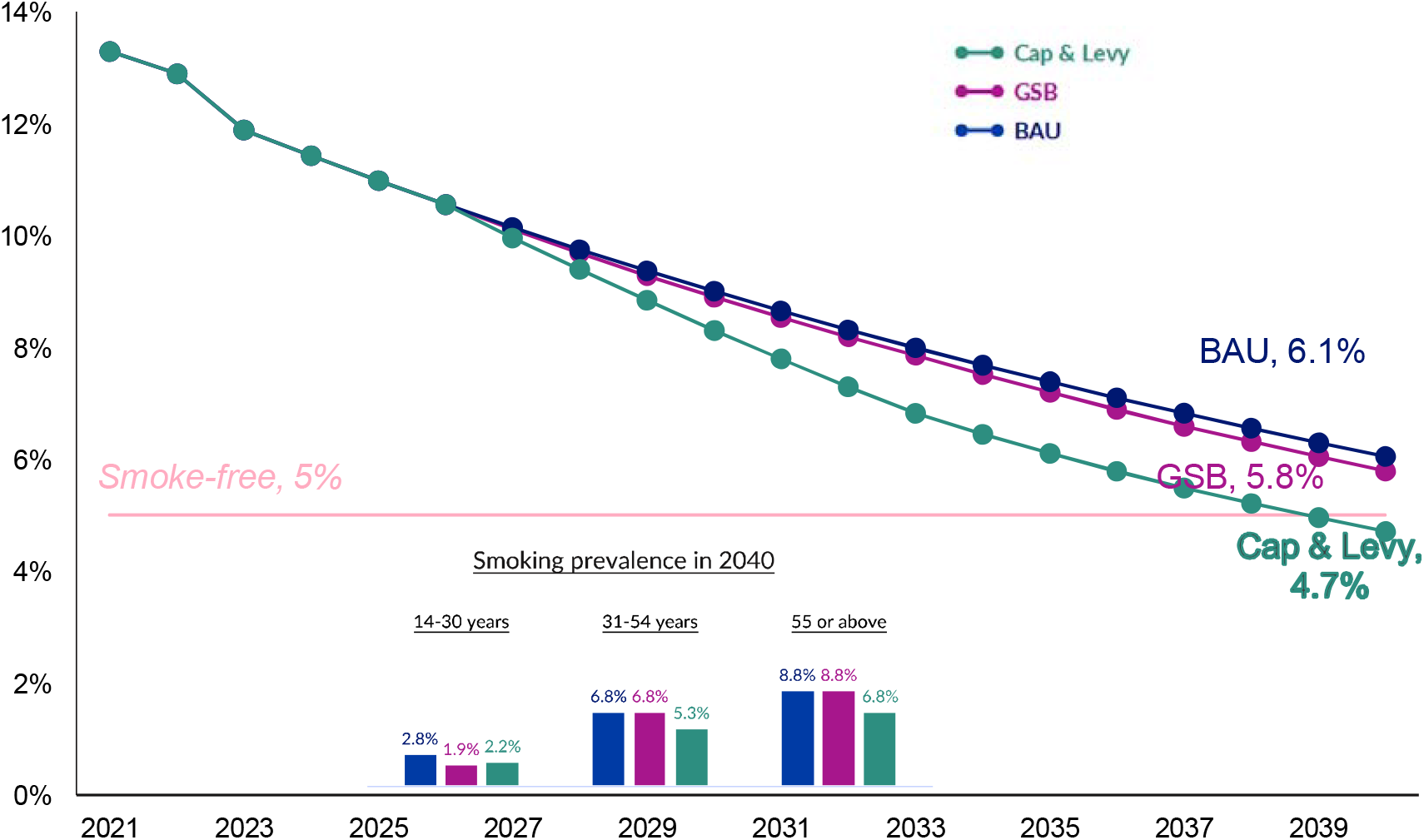
Projected impact of a Cap-and-Levy Scheme on smoking prevalence, 2023-2040. *Notes: Smoking prevalence is projected to fall below the 5% smoke-free threshold by 2039 under the cap-and-levy scheme*.

By 2040, tobacco sales would be halved -dropping to 3.7 billion stick equivalents compared to 7.5 billion under the GSB -and the prevalence of illicit trade is projected to be lower (1.1% vs. 1.4% under GSB). The cap-and-levy model also better supports transitions to reduced-risk products (RRPs) and is easier to enforce, focusing regulation on producers rather than consumer behaviour (similar to approaches in the EU ETS and UK Vehicle Emissions Trading Scheme; cf. European Commission, 2024).

Figure 6 presents detailed results of the Effectiveness scenario, showing a significant reduction in illicit smoking prevalence to 1.1% by 2040 under the Cap-and-Levy scheme, compared to 1.4% under GSB and 1.2% under BAU. This lower illicit prevalence can be attributed to the gradual reduction in supply, which allows consumers more time to adjust, and the focus on regulating producers, which simplifies compliance.

**Figure 6:**
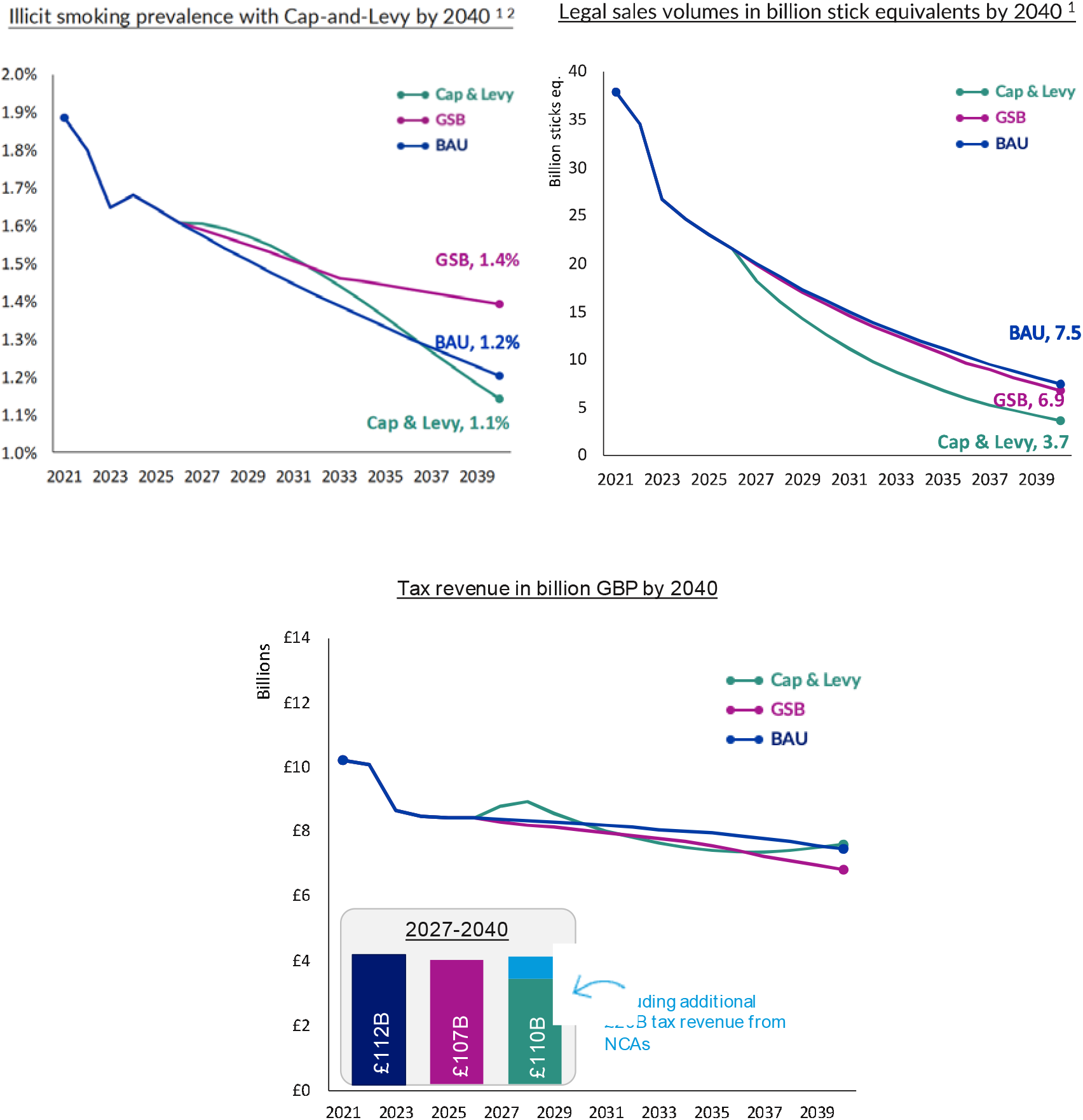
Illicit Smoking, Legal sales volume, and Tax Revenue, under Effectiveness Scenario.

Legal sales volumes decrease more rapidly under the Cap-and-Levy scheme in the Effectiveness scenario, reaching 3.7 billion stick equivalents by 2040, compared to 6.9 billion under GSB and 7.5 billion under BAU. This accelerated decline results from the supply cap affecting all age groups and the levy further raising prices.

However, this scenario leads to a reduction in tax revenue from combustible tobacco, with total revenue from 2027 to 2040 projected at £91.0 billion, compared to £107.4 billion under GSB and £112.3 billion under BAU. This loss is partially offset by an additional £19.5 billion in tax revenue from non-combustible alternatives, assuming 90% of smokers switch to these products.

To synthesize the findings of the scenario analysis, Table 2 provides a comparative summary of key outcomes across the Business-as-Usual (BAU), Generational Sales Ban (GSB), and Cap-and-Levy scenarios. This comprehensive comparison evaluates each policy’s impact on smoking prevalence, tax revenue, consumption volumes, and illicit market activity by 2040, alongside the primary assumptions underpinning each scenario.

**Table 2:**
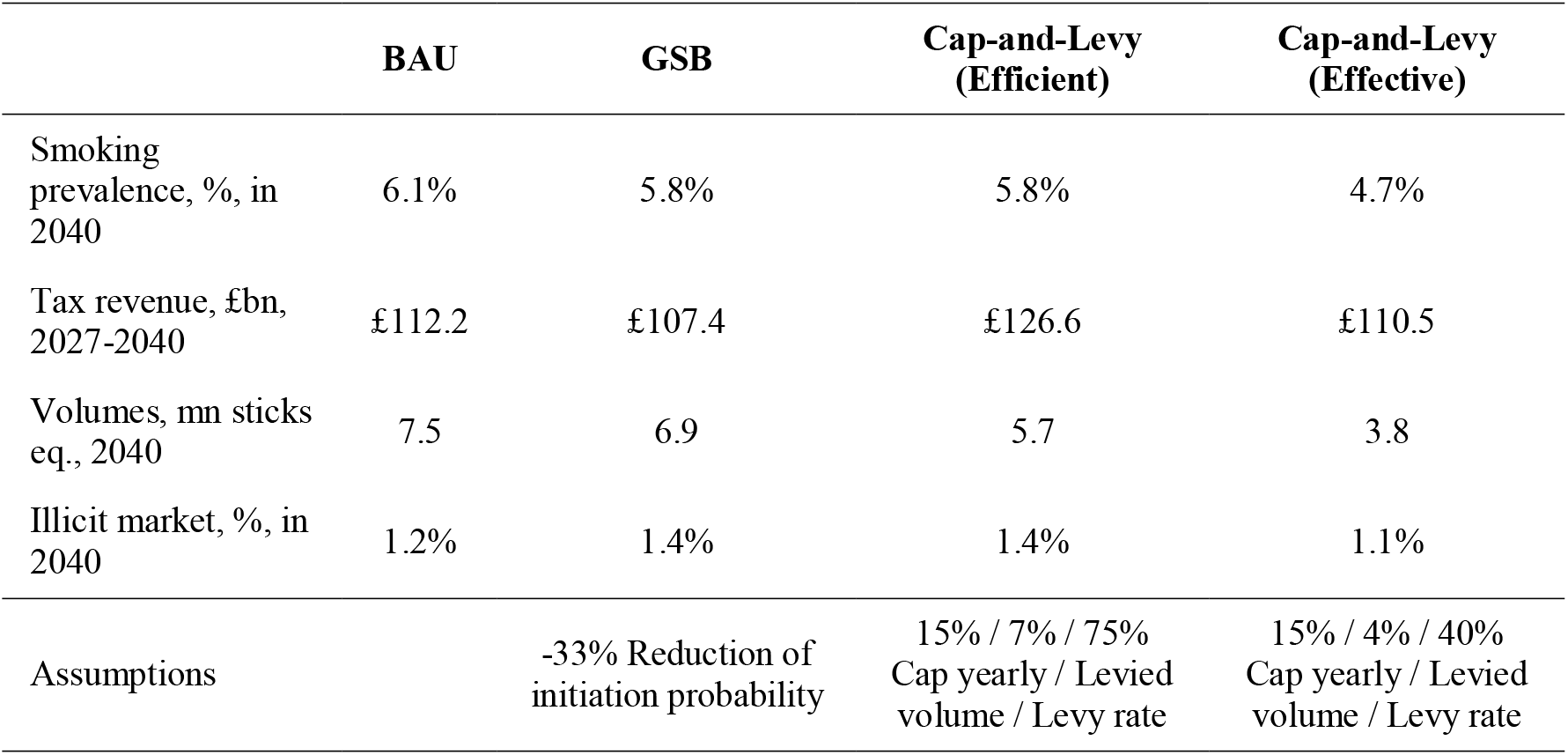
Summary of Result for each Policy and Scenario.

The table reveals that by 2040, smoking prevalence is projected to be lowest under the Cap-and-Levy (Effectiveness) scenario at 4.7%, compared to 5.8% under the Generational Sales Ban (GSB) and 6.1% under the Business-as-Usual (BAU) scenario. From a fiscal perspective, the Cap-and-Levy (Efficiency) scenario is projected to generate the highest tax revenue from 2027 to 2040 at £126.6 billion, exceeding both the £107.4 billion projected under the GSB and the £112.2 billion under the BAU scenario.

To wrap up, the results summarized in Table 2 demonstrate the significant advantages of the Cap-and-Levy scheme over the Generational Sales Ban (GSB). While both policies aim to reduce smoking prevalence, the Cap-and-Levy framework -particularly in its Effectiveness-Oriented configuration-projects a more substantial reduction in smoking rates by 2040. Furthermore, the Cap-and-Levy scheme shows a remarkable capacity to address the illicit tobacco market. As indicated in Table 2, the Effectiveness-Oriented scenario is the only policy that reduces illicit trade compared to the Business-as-Usual scenario, highlighting an additional benefit over the GSB, which is projected to increase illicit trade.

## Conclusion

The United Kingdom’s commitment to achieving a smoke-free society by 2030, defined by a smoking prevalence of less than 5%, represents a significant public health ambition (Department of Health and Social Care, 2019). Despite consistent declines in smoking rates over recent decades, the nation remains off track to meet this critical target. In response to this challenge, the government has put forward a novel policy proposal: a Generational Sales Ban (GSB). This legislation aims to prohibit the sale of tobacco products to individuals born after January 1, 2009. The rationale behind the GSB is to progressively eliminate smoking by preventing future generations from ever legally accessing tobacco.

While the GSB represents an innovative approach to tobacco control, this study posits that it suffers from inherent limitations that may hinder its effectiveness in achieving the 2030 smoke-free goal and could potentially generate unintended negative consequences. Primarily, the GSB focuses its intervention solely on future generations of smokers, thereby neglecting the significant health burden and economic costs associated with the current population of smokers (ONS, 2023). Data consistently reveals that smoking prevalence remains disproportionately high among individuals in lower socioeconomic strata, who often face greater obstacles in their cessation efforts (Hiscock et al., 2012; Reid et al., 2010). By overlooking this substantial segment of the smoking population, the GSB may only yield gradual reductions in overall smoking prevalence, potentially falling short of the ambitious 2030 target.

Furthermore, the implementation of the GSB carries a considerable risk of inadvertently fuelling the already substantial illicit tobacco market in the UK, which currently accounts for an estimated 26% of total tobacco consumption (HMRC, 2023). The creation of a legal distinction based on birth year could incentivize the development of black market channels to cater to those legally restricted from purchasing tobacco. Research on similar age-restriction policies, such as the study by Cotti et al. (2024), suggests that affected smokers may adapt their behaviour by seeking out sources in different jurisdictions rather than quitting altogether (Cotti et al., 2024). This could lead to an increase in the consumption of unregulated and potentially more harmful tobacco products, undermining public health objectives and further empowering criminal organizations. Additionally, a thriving illicit market would inevitably erode tax revenues generated from legal tobacco sales, impacting public finances.

Moreover, the evidence regarding the effectiveness of broad public smoking bans in reducing overall smoking prevalence is mixed. Studies, such as that by Jones et al. (2011), have indicated that such bans may have a limited impact on the total number of smokers (Jones et al., 2011). They can even lead to unintended consequences, such as the displacement of smoking to private spaces, as observed by Adda and Cornaglia (2010), without delivering significant public health gains in the short to medium term (Adda & Cornaglia, 2010). This suggests that solely restricting access in certain environments may not be sufficient to drive down overall consumption rates.

In contrast to the limitations of the GSB, this study advocates for the adoption of a comprehensive cap-and-levy scheme as a more effective and economically sound alternative for achieving the UK’s smoke-free ambitions. A cap-and-levy system operates by placing a progressively declining cap on the total volume of tobacco supplied into the market. Simultaneously, a levy or fee is imposed on tobacco manufacturers and importers for each unit of tobacco they introduce. This dual mechanism directly addresses both the supply and demand sides of tobacco consumption. The declining cap ensures a gradual but consistent reduction in the availability of tobacco products, making it increasingly difficult for individuals to access them over time. The levy, by increasing the cost of tobacco products, incentivizes smokers to reduce their consumption or quit altogether, while also generating substantial revenue that can be reinvested in public health initiatives, including smoking cessation programs and awareness campaigns.

The implementation of a cap-and-levy scheme offers several key advantages over the GSB. Firstly, it exerts a direct influence on the entire smoking population, including both current and future smokers. The progressively decreasing supply and the increasing cost of tobacco will encourage cessation among existing smokers and deter initiation among younger individuals, regardless of their birth year. This broader impact is crucial for achieving the ambitious 2030 smoke-free target in a timely manner. Secondly, by directly controlling the supply chain, a cap-and-levy system can effectively curtail the growth of the illicit tobacco market. As the legal supply diminishes, the economic incentives for illicit trade are reduced, as the overall availability of tobacco products decreases. Furthermore, the revenue generated by the levy can be used to strengthen enforcement measures against illicit trade, further bolstering its effectiveness. Thirdly, the revenue generated by the levy provides a sustainable funding stream for vital public health programs aimed at reducing smoking prevalence and treating smoking-related illnesses. This reinvestment of funds directly addresses the health consequences of smoking and supports individuals in their efforts to quit.

Modelling projections based on the principles of supply and demand elasticity, as explored by Pryce et al. (2024) and the World Bank (1999), indicate that a well-designed cap-and-levy scheme has the potential to achieve the UK’s smoke-free target by approximately 2039. This projection takes into account the likely behavioural responses of smokers to increasing prices and decreasing availability. Furthermore, the analysis suggests that such a scheme could lead to a significant increase in tax revenue compared to the GSB, which may lead to a decline in tax receipts due to the potential growth of the illicit market. This additional revenue can be strategically allocated to fund public health initiatives and offset the healthcare costs associated with smoking-related diseases. Moreover, the cap-and-levy approach is projected to substantially reduce the prevalence of illicit trade by making it more difficult and less profitable for illegal operators to function effectively (Prieger & Kulick, 2016).

The policy implications of this research are significant. The findings strongly suggest a need for policymakers to reconsider the current singular focus on the Generational Sales Ban and to give serious consideration to the adoption of a cap-and-levy framework as a more comprehensive and economically sound tobacco control strategy. Implementing a cap-and-levy scheme would represent a paradigm shift in the UK’s approach to tobacco control, moving from a primarily age-based restriction to a market-based mechanism that directly addresses consumption and supply. This shift has the potential to accelerate progress towards a smoke-free UK, ensuring greater fiscal sustainability and minimizing the unintended negative consequences associated with the GSB. To effectively implement a cap-and-levy system, policymakers would need to establish a robust regulatory framework for monitoring and enforcing the cap on tobacco supply and collecting the levy. Collaboration with tobacco manufacturers and retailers would be essential to ensure a smooth transition and minimize disruption to the legal market. Public communication campaigns would also be crucial to educate smokers about the rationale behind the policy and to promote the availability of cessation support services.

In conclusion, while the UK’s ambition to achieve a smoke-free status by 2030 is commendable, the proposed Generational Sales Ban, despite its novelty, presents significant limitations in its scope and potential for unintended negative consequences, including fuelling the illicit market and reducing tax revenues (HMRC, 2023; ONS, 2023; Snowdon, 2024). Furthermore, the availability of appealing and accessible alternatives to combustible tobacco is a crucial factor in mitigating the risk of an expanding illicit market under restrictive policies. This study provides compelling evidence that a cap-and-levy scheme offers a more comprehensive and economically viable alternative (World Bank, 1999). By directly addressing both the supply and demand of tobacco, this approach is projected to achieve the smoke-free target by 2039, increase government tax revenue, and significantly reduce the prevalence of illicit trade (Prieger & Kulick, 2016). The policy implications of this research underscore the urgent need to reconsider the current tobacco control strategy and to adopt a cap-and-levy framework to accelerate progress towards a smoke-free UK, while simultaneously ensuring long-term fiscal sustainability and minimizing the detrimental unintended consequences associated with a generational sales ban.

## Data Availability

Data is publicly available.

## Notes

### Competing Interest Statement

The authors have declared no competing interest.

### Funding Statement

This study did not receive any funding.

